# Therapeutic effectiveness of interferon alpha 2b treatment for COVID-19 patient recovery

**DOI:** 10.1101/2020.07.28.20157974

**Authors:** R Pereda, D González, HB Rivero, JC Rivero, A Pérez, LR López, N Mezquia, R Venegas, Julio Raul Betancourt, RE Domínguez

**Affiliations:** Intensive Medicine Department, Medical College of Havana. 23 Street 202, Vedado, Plaza de la Revolución, ZIP code 10400. Havana, Cuba; Internal Medicine Department, Pedro Kouri Institute. Avenida Novia del Mediodía, KM 6 ½, La Lisa, ZIP code 11400, Havana, Cuba; Internal Medicine Department, Enrique Cabrera General Hospital. Calzada de Aldabó 11117, Altahabana, Arroyo Naranjo, ZIP code 10800, Havana, Cuba; Intensive Medicine Department, Miguel Enríquez Surgical Clinical Hospital. Ramón Pinto 202, 10 de octubre, ZIP code 10700, Havana, Cuba; Intensive Medicine Department, Enrique Cabrera General Hospital. Calzada de Aldabó 11117 Corner E, Altahabana, Arroyo Naranjo, ZIP code 10800, Havana, Cuba; Intensive Medicine Department, Juan Manuel Marquez Pediatric Hospital. Avenida 31, Marianao, ZIP code 11400, Havana, Cuba; Intensive Medicine Department, Luis Díaz Soto Central Military Hospital. Ave Monumental km 2½ Eastern Havana, ZIP code 19130, Havana, Cuba; Intensive Medicine Department, Manuel Piti Fajardo Surgical Clinical Military Hospital. New Abel Santa María, Santa Clara, ZIP code 50100, Villa Clara, Cuba; Intensive Medicine Department, Octavio de la Concepcion y la Pedraja Military Hospital. Cornelio Porro Street 52, Camagüey, ZIP code 70200, Camagüey, Cuba

**Keywords:** interferon, COVID-19, SARS-Cov-2

## Abstract

**Background:** Effective antiviral treatments are required to contain the ongoing coronavirus disease 2019 (COVID-19) pandemic. A previous report in 814 patients COVID-19 positive in Cuba provided preliminary therapeutic efficacy evidence with interferon-α2b (IFN-α2b) from March 11 to April 14, 2020. This study, re-evaluates the contribution of IFN-α2b on the evolution of all patients, after 98 days of the epidemic, in a period from March 11 to June 17, 2020.

**Method:** A prospective observational study was implemented to monitor a therapeutic intervention with IFN-α2b used in the national protocol for COVID-19 attending in Cuba. Were included patients with positive throat swab specimens by real time RT-PCR who gave informed consent and had no contraindications for IFN treatment. Patients received therapy as per the Cuban COVID protocol that included a combination of oral antivirals (lopinavir/ritonavir and chloroquine) with intramuscular or subcutaneous administration of IFN-α2b The primary endpoint was the proportion of patients discharged from hospital, secondary was the case fatality rate and several outcomes related to time variables were also evaluated.

**Results:** From March 11^th^ until June 17^th^, 2295 patients had been confirmed SARS-CoV-2 positive in Cuba, 2165 were treated with Heberon^®^ Alpha R and 130 received the approved protocol without IFN. The proportion of fully recovered patients was higher in the IFN-treated compared with non-IFN treated group. Prior IFN treatment decreases the likelihood of intensive care and increases the survival after severe or critical diseases. The benefits of IFN were significantly supported by time variables analyzed.

**Conclusions:** This second report confirm the preliminary evidences from first for the therapeutic effectiveness of IFNα-2b for SARS-Cov-2 infection and postulated that Heberon^®^ Alpha R is the main component within the antiviral triad used as a therapeutic intervention in the Cuban protocol COVID-19.

## INTRODUCTION

The coronavirus disease 2019 (COVID-19) emerged in China in December 2019^1^. The responsible virus, severe acute respiratory syndrome coronavirus 2 (SARS-CoV-2), belongs to a distinct classification from the human severe acute respiratory syndrome CoV (SARS-CoV) and Middle East respiratory syndrome CoV (MERS-CoV)^2^. The clinical spectrum of COVID-19 varies from asymptomatic infection or mild symptoms to severe acute respiratory illness and death^3^. According to the COVID-19 situation report issued by World Health Organization (WHO) on 11^th^ July 2020, COVID-19 pandemic affects more than 213 countries around the world with more than 12 million cases have been diagnosed and over 556 thousand patients have died^4^. Effective antiviral treatments are required to contain the epidemics; several candidates are already being investigated, including type 1 interferon (IFN-α/β)^5^.

At present, a RNA-dependent RNA polymerases (RdRp), remdesivir, is the first medicine against covid-19 approved in the European Union, mainly based on preliminary data from a trial published in the New England Journal of Medicine and limited to patients with severe disease^6^. Remdesivir has also been approved for emergency use in severely ill patients in the United States (US), India, and South Korea and has received full approval in Japan^7^. Favipiravir^8^, an anti-influenza medication that also is an RdRp inhibitor, was approved for medical use in Russia using a generic version named Avifavir.

IFN-α, a member of the first line of natural antiviral defense activates the innate immune response against the virus and the mechanism of inhibition of viral replication^9^. After COVID-19 outbreak, a strong anti-SARS-CoV-2 activity in cultured cells has been reported for IFN-α, demonstrating its therapeutic potency for COVID-19^10^. Guidelines issued by the expert committee of WHO identified IFN-α2b as a potential antiviral for the treatment and prevention of COVID-19^11^. Early on in the outbreak, the Chinese government recommended the use of IFN-α for the treatment of COVID-19^12^. Several ongoing clinical studies evaluating IFN-α2b for the treatment of COVID-19 are registered at clinicaltrials.gov^13^ and already provided clinical evidences for therapeutic efficacy^14^.

Heberon^®^ Alpha R is a human recombinant IFN-α2b formulation produced by the Center for Genetic Engineering and Biotechnology, Havana, Cuba, with demonstrated antiviral efficacy and a proven safety profile over 34 years^15^ and is one of the drugs used in the Cuban protocol^16^ for the management of COVID-19. A prospective observational study was implemented to monitor therapeutic interventions used in the national protocol for COVID-19 attending in Cuba. The preliminary results in 814 patients included from March 11 to April 14, 2020 was reported previously^17^ and here we reanalyze the evolution of all patients COVID-19 positive in Cuba and report the efficacy of IFN-α2b after 98 days of the epidemic, a period from March 11 to June 17, 2020.

## MATERIALS AND METHODS

### Patients

Two groups of individuals were admitted to the hospital, according to the case classification criteria defined in the Cuban protocol: 1) people with suspected COVID-19 due to clinical respiratory symptoms, such as fever, fatigue, cough, headache, shortness of breath and nasal discharge in the last 14 days; 2) subjects who had contact with a patient with confirmed or suspected COVID-19 infection and were followed up for 14 days for possible infection. Each patient had to declare their date of symptom onset. Persons, with or without symptoms, were considered COVID-19 confirmed based on SARS-CoV-2 detected by real time polymerase chain reaction (RT-PCR).

### Treatment

Individuals below age 50 without comorbidities and suspected COVID-19 started treatment at day of hospital admission. A combination of oseltamivir 75 mg every 12 hours for five days, azithromycin 500 mg daily for 3 days and IFN-α2b human recombinant (Heberon^®^ Alpha R) by intramuscular injection 3 million IU 3 times per week, for 4 weeks were prescribed. Once individuals were COVID-19 confirmed patients, lopinavir/ritonavir (LPV/RTV) (250 mg, one capsule b.i.d. (500 mg/day) for 30 days and chloroquine (CQ) 150 mg, one tablet twice a day (300 mg/day) for 10 days were introduced and oseltamivir and azithromycin leave off. People with suspected COVID-19 in age 50 or older with or without comorbidities and subjects below age 50 with comorbidities were considered high-risk patient and antiviral treatment initiated directly with LPV/RTV + CQ + IFN-α2b. The asymptomatic close contact subjects began treatment within 48 hours after a positive RT-PCR confirming SARS-CoV-2 infection and received LPV/RTV + CQ + IFN-α2b. To treat pediatric cases, all drugs were adjusted for age and weight or body surface (i.e. IFN: 100.000 IU/Kg) and use subcutaneous route for IFN-α2b.

Individuals themselves, parents or legal representative had to give informed consent and have no contraindications for treatment with IFN described in the product information sheet, to receive Heberon^®^ Alpha R as approved in the Cuban protocol for COVID-19. Patients in severe COVID-19, with contraindications or who did not consent to receive IFN were treated with the Cuban protocol lacking IFN, i.e. only LPV/RTV and CQ. For those cases in treatment ongoing whose disease progressed to become severe and critical, requiring intensive care unit (ICU) support, treatment with Heberon^®^ Alpha R was promptly stopped.

Addition of Heberon^®^ Alpha R to the Cuban protocol was approved by the Cuban National Regulatory Authority which maintained surveillance of the scientific evidence obtained. The protocol of this study was evaluated and approved by a centralized Research Ethics Committee, representing all hospital institutions enrolled in the diagnosis and treatment of patients. This clinical research is registered with the code RPCEC00000318 in the Cuban Public Registry of Clinical Trials^18^. Data presented are from patient medical records provided by the Cuban Ministry of Public Health, in accordance with the policies established for an investigation in a pandemic scenario.

### PCR confirmation of SARS-CoV-2

A qualitative RT-PCR for SARS-CoV-2 was performed. Throat swab specimens from the upper respiratory tract of patients were placed into collection tubes prefilled with 150 μL of virus preservation solution, and total RNA was extracted using commercial kits: LightMix® Modular Sarbecovirus E-gene (Roche), LightMix® Modular SARS-COV-2 (COVID19 RDRP-GENE) (Roche), LightMix® Modular EAV RNA Extraction Control (Roche), LightCycler Multiplex RNA Virus Master (Roche), QIAamp® Viral RNA MiniKit (250) (Quiagen). Two consecutive RT-PCR positive results, at least 24 hours apart, was required before the start of treatment. Evolutionary PCR was performed 14 days later (at the end of treatment) and repeated weekly, if necessary, to undetectable values. Samples were designated positive (+) or negative (-).

Hematological and biochemical profiles were assessed at admission and every 48 hours using routine clinical laboratory procedures. Chest radiological evaluation and cardiovascular function monitoring by electrocardiogram was made before treatment and daily after started it.

## Endpoints

### Main outcomes

The primary endpoint was the proportion of patients discharged from hospital (i.e. discharge criteria were according to clinical, radiological and laboratory evaluations). Clinical criteria: patient in stable condition and afebrile for more than 3 days, regular breathing and normal respiratory rate, clear conscience, unaffected speech and normal diet. Radiological criteria: significant improvement without signs of organ dysfunction in lung images. Laboratory criteria: Two consecutive PCR (-) with at least 24 hours apart. The secondary endpoint was the case fatality rate, (CFR), defined as the number of confirmed deaths divided by the number of confirmed cases.

### Others outcomes related to time variables

The time elapsed to the hospital discharged was an endpoint for all patients. A threshold time definition was according the patient clinical classification: for symptomatic subject was defined as the number of days elapsed from the onset of symptoms up to meet discharged criteria; for asymptomatic the timeline began on hospital day admission. The full recovery time after COVID-19 diagnostic by PCR (+) was also endpoint for all cases. Date of confirmed PCR (+) was considered as date of onset of COVID-19 and the need of treatment, an appropriate time to match point the disease course between all patients. Endpoints applied to cohort requiring ICU were the days from symptom onset and hospital admittance to ICU admission and the time to death from these three timelines.

### Statistical analysis

Descriptive statistics (Tables, group means and interquartile range reported in the text) convey the data as-is. The endpoints were adjusted by age, sex, and the presence of comorbidities. Note that age was considered at every 10-year age interval from older than 19 years. From contingency tables chi-square calculator tests and Fisher exact test calculator for association between two categorical variables were used. The odds ratio (OR) was applied as a measure of association between treatments and outcomes. The non-parametric Mann-Whitney U test was used for comparing independents samples

## RESULTS

### Patients included and treatment received

The Cuban protocol^17^ for clinical management of COVID-19 was applied since March 11^th^, 2020. By June 17th, according to the Cuban Ministry of Public Health^19^, 2295 individuals had been confirmed positive for SARS-CoV-2 infection and included in this observational study, 2053 of them (89.5%) adults and 242 (10.5%) in pediatric age. On hospital admission 1234 (53.8%) individuals were asymptomatic.

To begin treatment, two clinical forms of uncomplicated disease were predominated: 1) minimally symptomatic disease according to nonspecific signs such as fever, cough, sore throat, nasal congestion, slight headache and malaise, without dehydration, dyspnea, or sepsis evidences; 2) mild pneumonia, due to the presence of the above symptoms and polypnea with partial oxygen saturation (SpO_2_) above 90%, but without signs of respiratory failure or severity.

In 44 (1.9%) patients a severe pneumonia was detected before start treatment and their hospital admission was directly to ICU due to positive clinical symptoms, chest radiological imaging and respiratory rate over 30 breaths / min, limitation of thoracic expandability, central cyanosis, SpO_2_ below 90% and pleuritic pain. Acute respiratory distress syndrome (ARDS) associated was found in 17 of these (38.6%) patients.

**Table 1** summarizes the patient demographics and clinical characteristics of COVID-19 cases evaluated in this observational study. IFN treatment was prescribed in 2165 (94.3%) patients and 130 (5.7%) received the Cuban protocol without IFN, 124 due to contraindications and six declined informed consent. The population and treatment received were dependent variables (*p*=0.002), 1948 (94.9%) adults and 217 (89.7%) children were treated with IFN. All IFN-treated patients received also LPV/RTV, 11 of them (0.5%) started LPV/RTV a few days after IFN treatment had been initiated. A triple therapy IFN-α2b, LPV/RTV and CQ was applied in 2105 (91.7%) patients

**Table 1.** Demographics & clinical characteristics of patient cohort.

|  | IFN<br>n=2165 | No IFN<br>n=130 | *p* value |
| --- | --- | --- | --- |
| Age<sup>a</sup>, years | 44 (**0-100**) | 68 (**0-101**) | $7.0 \times 10^{-14}$ |
| Male (%) | 1101 (50.9%) | 68 (52.3%) | 0.817 |
| Co-morbidities<sup>b</sup> | 237 (10.9%) | 102 (78.5%) | $1.0 \times 10^{-5}$ |
| Asymptomatic disease (%) | 1211 (55.9%) | 23 (17.7%) | $1.0 \times 10^{-5}$ |

<sup>a</sup> median and range is reported
<sup>b</sup> high blood pressure, diabetes mellitus, ischemic heart disease, chronic kidney failure, asthma, obesity, chronic obstructive pulmonary disease, malignancy, liver cirrhosis, gastritis, hypothyroidism, glaucoma, myasthenia gravis, HIV.

### Patient demographics and clinical characteristics

The median age of all cases was 45 years (range 19 days to 101 years; interquartile range, 28– 58 years) with a non-significant difference between the sexes (*p*=0.511). In the distribution of patients by sex, the numbers of men (*p*=0.006) at ages 61-70 and women (*p*=0.035) above age 90 were significantly higher. Both (*p*=0.445) women and men peaks COVID-19 incidence at ages 51-60. Comorbidities were present in 339 (14.8%) patients with a similar incidence between the sexes (p=0.923) and significant higher in those over age 51 not treated with IFN. The main underlying diseases were high blood pressure (7.9%), diabetes mellitus (4.2%) and ischemic heart disease (3.2%).

### Proportion of patients discharged from hospital

On June 17, 187 IFN-treated patients and one IFN-non treated remained hospitalized in stable health condition under the care protocol (currently considered as patients with unknown outcome), while 2022 (88.1%) had recovered from COVID-19 and had been discharged from the hospital. Of the individuals with known results, the highest proportion that became PCR (-) for SARS-CoV-2 and that resolved their disease were those treated with IFN (1958; 99.0% vs. 64; 49.6%, *p*=1.0 × 10^−5^). According to the Odds Ratio estimation, an individual treated with Heberon^®^ Alpha R had at least, 56.8 times greater likelihood to achieve recovery.

### Effects of IFN treatment on CFR

On the cut-off date analyzed in this report, the Cuban Overall CFR was 3.70% and 0.92% for patients treated with IFN alpha-2b. Both data were lower than those reported at the same time by WHO^20^ (Global CFR=5.45%) and Pan American Health Organization^21^ (PAHO CFR=5.24% for the region of the Americas). Clinical, radiological and virological evaluation revealed that there were more patients not treated with IFN-α2b who had an unfavorable outcome of disease (i.e. presence of clinical and pulmonary symptoms and/or detectable virus).

### CFR in patients with COVID-19 in severe or critical stage

To better address the influence of IFN treatment on CFR, we considered the clinical status of patients in the context of severity of disease, using admission to ICU as an appropriate time point to match the criterion for worsened disease. During the course of treatment, 180 patients (7.8% of all the COVID-19 cases) were admitted to the ICU (**Table 2**). Notably, only 77 had been treated with IFN, representing 3.6% of the total number of this treatment cohort in the study. IFN treatment was withdrawn for those patients who progressed to severe or critical disease. Nevertheless, the CFR in presence of ARDS was lower in IFN previously treated individuals (10; 47.6% vs. 22; 84.6%, *p*=0.02). The effects of IFN treatment on subsequent CFR revealed that prior IFN treatment was associated with a 64.0 times greater likelihood of not requiring ICU and increases 2.6 times the probability of survival after admission to ICU. The benefits of IFN were significantly supported by the results of all the time variables analyzed (**Table 3**).

**Table 2.** Patients with ICU requirement.

|  | IFN<br>n=2165 | No IFN<br>n=130 | *p* value |
| --- | --- | --- | --- |
| Serious disease (%) | 42 (1.9%) | 48 (36.9%) | $< 1.0 \times 10^{-5}$ |
| Critical disease (%) | 35 (1.6%) | 55 (42.3%) | $< 1.0 \times 10^{-5}$ |
| ARDS (%) | 21 (1.0%) | 26 (20.0%) | $< 1.0 \times 10^{-5}$ |
| CFR<sup>a</sup> | 20 (26.0%) | 65 (63.1%) | $< 1.0 \times 10^{-5}$ |
| Age of deceased<sup>b</sup> | 67.5 (49-91) | 77.0 (35-101) | 0.150 |

<sup>a</sup> regarding only ICU patients
<sup>b</sup> median is reported

**Table 3.** Effects of IFN treatment on CFR expressed in time variables.

| Times intervals |  | Treatment<sup>a</sup> |  |  |
| --- | --- | --- | --- | --- |
| T1 | T2 | IFN | No IFN | *p* |
| Symptom onset | ICU admission | 10.0<br>[6.0, 13.0] | 6.0<br>[3.0, 10.0] | 0.003 |
| Hospital admission | ICU admission | 6.0<br>[2.0, 10.0] | 2.0<br>[0.0, 5.0] | 0.007 |
| Symptom onset | Death | 16.0<br>[12.0, 21.0] | 9.0<br>[5.0, 14.0] | 1.0 x 10<sup>-5</sup> |
| Hospital admission | Death | 13.0<br>[6.0, 17.0] | 5.0<br>[2.0, 10.0] | 0.002 |
| ICU admission | Death | 7.0<br>[5.0, 11.0] | 2.0<br>[0.0, 4.0] | 1.0 x 10<sup>-5</sup> |

<sup>a</sup> Median in days and interquartile range [Q1, Q3] is reported

A detailed view in the health status of patients at the time of hospital admission revealed that 10 subjects death immediately without received none medication and later COVID-19 confirmation. After CFR correction analysis done without considered these patients, the IFN cohort maintained a significant outcome compared to IFN non-treated individuals (*p*=3.0 × 10^−5^).

### Effects of age, co-morbidities, sex and symptomatic disease on CFR

Patients died from COVID-19 in Cuba were adults. Thereby, the analysis related to age and lethality was made at every 10-year age interval from older than 19 years. Ages 51-60 were the only ones lacking significant association to CFR. There were 47 deceases in ages 71-90, 55.3% from overall. Deaths predominated in male (*p*=0.042), patients with comorbidities (*p*=1.0 × 10^−5^) and symptomatic individuals (*p*=1.0 × 10^−5^). In male over 51 years, arterial hypertension represented the major risk predictors for death.

IFN-related CFR was affected by ages 61-70 (*p*=0.007) and ages 81-90 (*p*=2.0 × 10^−4^). However, the effects of IFN treatment remain significant regardless of age interval compared to those not treated with IFN (p-values ranged 1.5 × 10^−2^ to 1.0 × 10^−5^). Although the IFN-related CFR was affected by comorbidities (*p*=1.0 × 10^−5^), did not negate or eliminate the difference in CFR between IFN treated individuals and those not treated with IFN (*p* =1.0 × 10^−5^). No association was found between sex and lethality in those treated with IFN (*p*=0.138).

## Discussion

Our previous report in 814 patients COVID-19 positive in Cuba provided preliminary therapeutic efficacy evidence with IFN-α2b from March 11 to April 14, 2020^17^. In this study we increase the sample size extending the analysis up to June 17, 2020, representing 98 days of COVID-19 epidemic in the country and a re-evaluating of IFN-α2b contribution on the evolution of all confirmed patients was realize. The results obtained on rates of recovery and case fatalities related to IFN-treated patients endorsed our previous findings showing the potential efficacy of human IFN-α2b in suppressing SARS-CoV-2 infection and reducing progression to severe disease in 814 patients^17^. Like in the earlier analysis at the first month of the epidemic in Cuba, but now in a more numerous population, the data over 98 days reveals a greater proportion of patients recovered after treatment with IFN-α2b. This consistent pattern should be considered strong evidence that intramuscular administration of this product is beneficial, useful and effective to achieve therapeutic effect in patients with COVID-19. The survival IFN-related achieved for severe and critical patients postulated that Heberon^®^ Alpha R is the main component within the antiviral triad used as a therapeutic intervention in the Cuban protocol COVID-19.

Ours two reports on the intramuscular administration for IFN-α2b contrast with the earlier published findings of inhaled IFN-α2b^14^. An aerosol administration has the advantage of specifically targeting the respiratory tract; however, the pharmacodynamics and pharmacokinetics of this mode of administration are not known^9^. Conversely, intramuscular and subcutaneous administration are well described for Heberon^®^ Alpha R and their safety profiles has been extensively studied by pharmacodynamics and pharmacokinetics and have already proven safe in a considerable number of clinical trials^16^.

The key explaining the large recovery of patients after used IFN-α2b in our data is a therapeutic strategy based on recommendation by Sallard et al^6^ for IFN-I administration in the early phases of the infection to optimize antiviral therapy. Morover, a recently news published in Science mentions at least five studies since April 2020 have found that interferon treatment has a protective effect in cells and in mice infected with SARS-CoV-2^22^. After our experience, we postulate that IFN treatment applied promptly to onset symptoms increases significantly the therapeutic benefit to COVID-19 cases. The active screening in close contact asymptomatic individual was also vital for made a positive difference in the evolution of the disease with Heberon^®^ Alpha R prescription early in infection.

The intramuscular or subcutaneous route for IFN-α2b administration and its demonstrated efficacy on early phases of virus infection, even in asymptomatic patients represent two important advantages compared to intravenous remdesivir regimen limited to severe patients. The cellular signaling system of IFN alpha let to blocking the synthesis of new viral particles and at the same time lead to the destruction of existing ones. This wide antiviral effect is restricted for traditional antiviral drugs, intended only to inhibit viral replication. The IFN alpha-2b exogenous administration causes a positive feedback mechanism by stimulating the production of endogenous IFN able to continuing the cellular signal leading to the biological effect of the IFN, even when it has disappeared from the blood circulation. As a result, a sustained therapeutic response is expressed clinically thought a long-term viral clearance after treatment. Neither of these two favorable events is possible with generic antiviral drugs like favipiravir, whose depend on permanent level in blood for achieve therapeutic activity.

Demographic distribution of patients included in this study conformed to findings in China^23^, in several European^24^ and Latin-American^25^ countries and the US^26^ reporting a slightly higher number for male confirmed and CFR compared to females. Older persons with pre-existing hypertension and/or diabetes were more prevalent in our no-IFN treated cohort and this age-related trend of co-morbidities was similar to data reported by Zhou F et al^14^.

The proportion (10%) of confirmed COVID-19 children in Cuba seems notably when pediatrics reviews show that pediatric population have so far accounted for 1%-5% of diagnosed COVID-19 cases around globe^27^ The above can be explained by the methodology implemented by the Cuban protocol in which an early epidemiological screening was carried out independently of the presence of clinical symptoms and most of the patients were admitted to hospital and SARS-CoV-2 confirmed in the asymptomatic phase.

Data analyses in this study were limited, because the study includes unbalanced demographics between treatment arms of unequal size and endpoint outcomes, namely hospital discharge (recovery) and CFR for a large number of the cases remained unknown at the time of study termination. Regardless of the identified limitations, this report provides evidence of the effectiveness of IFN-α2b as an antiviral treatment for SARS-CoV-2 infection and suggests that the use of Heberon^®^ Alpha R at the doses and therapeutic regimen employed may contribute to recovery from COVID-19. Furthers studies are needed for additional efficacy and safety profile evaluation of Heberon^®^ Alpha R, specifically a randomized clinical trial for comparison with other potential antivirals.

## Contributors

DG, RV, BJR and DRE were responsible for patient care and treatment, RHB, RJC, OA, LLR and MN made clinical oversight and clinical data collection, PR led the working group, analyzed and conducted data analysis, data interpretation, literature searches and manuscript writing.

## Data Availability

Data presented are from patient medical records provided by the Cuban Ministry of Public Health, in accordance with the policies established for an investigation in a pandemic scenario

